# Association of SARS-CoV-2 BA.4/BA.5 Omicron lineages with immune escape and clinical outcome

**DOI:** 10.1101/2022.07.31.22278258

**Authors:** Joseph A. Lewnard, Vennis Hong, Jeniffer S. Kim, Sally F. Shaw, Bruno Lewin, Harpreet Takhar, Sara Y. Tartof

**Author notes:** Addresses for correspondence: Joseph A. Lewnard, 2121 Berkeley Way, Berkeley, California 94720, Sara Y. Tartof, 100 South Los Robles, Pasadena, California 91101.

## Abstract

Expansion of the SARS-CoV-2 BA.4 and BA.5 Omicron subvariants in populations with prevalent immunity from prior infection and vaccination, and associated burden of severe COVID-19, has raised concerns about epidemiologic characteristics of these lineages including their association with immune escape or severe clinical outcomes. Here we show that BA.4/BA.5 cases had 15% (95% confidence interval: 9-21%) and 38% (27-49%) higher adjusted odds of having received 3 and ≥4 COVID-19 vaccine doses, respectively, than time-matched BA.2 cases, as well as 55% (43-69%) higher adjusted odds of prior documented infection. However, after adjusting for differences in epidemiologic characteristics among cases with each lineage, BA.4/BA.5 infection was not associated with differential risk of emergency department presentation, hospital admission, or intensive care unit admission following an initial outpatient diagnosis. This finding held in sensitivity analyses correcting for potential exposure misclassification resulting from unascertained prior infections. Our results demonstrate that the reduced severity associated with prior (BA.1 and BA.2) Omicron lineages, relative to the Delta variant, has persisted with BA.4/BA.5, despite the association of BA.4/BA.5 with increased risk of breakthrough infection among previously vaccinated or infected individuals.

---

The SARS-CoV-2 Omicron (B.1.1.529) variant emerged in late 2021 and rapidly achieved global dissemination, accounting for a majority of incident SARS-CoV-2 infections within the United States by late December, 2021.^1,2^ As of February, 2022, 58% of US adults and 75% of US children aged ≤17 years were estimated to have acquired SARS-CoV-2 infection, with nearly half of these infections occurring during the initial expansion of the BA.1 subvariant lineage.^3^ COVID-19 vaccination and naturally-acquired immunity from infection with pre-Omicron variants have generally been found to confer robust protection against clinically severe disease involving the BA.1 lineage, although at weaker levels when compared with protection against pre-Omicron variants.^4–7^ Thus, whereas expansion of the Omicron variant was associated with surges in COVID-19 hospitalizations and deaths, the proportion of Omicron cases resulting in severe illness has been lower than that experienced with prior variants and during periods with lower population-level immunity^4,8^

Following the initial peak in BA.1 infections within the US from December, 2021 to February, 2022, multiple Omicron lineages have driven subsequent surges in cases, leading to persisting clinical burden and extended timetables for implementation of COVID-19 mitigation measures. Although not associated with enhanced severity or risk of breakthrough infection after vaccination or natural infection,^4,9^ the BA.2 lineage surpassed BA.1 in incident cases within the US beginning in March, 2022. Subsequently, the BA.4 and BA.5 lineages have become dominant globally.^10^ BA.4 and BA.5 share a spike (S) protein harboring numerous mutations relative to BA.2, which may compromise the effectiveness of immune responses induced by prior infection and vaccination.^11^ Other mutations specific to BA.4 and BA.5 alter binding to human angiotensin-converting enzyme-2 and non-S antibodies derived from prior infection.^12,13^ However, clinical implications of the emergence of BA.4/BA.5 remain uncertain, as the burden of hospitalized and fatal COVID-19 cases observed during BA.4/BA.5 waves has varied widely across settings.^14^

Monitoring the relative severity of infections caused by successive SARS-CoV-2 lineages, and their capacity to evade vaccine- or infection-derived immunity, is of key importance to informing public health mitigation measures as SARS-CoV-2 establishes endemic circulation. We therefore compared clinical outcomes and characteristics of contemporaneous cases with BA.2 and BA.4/BA.5 lineage Omicron variant infections within the Kaiser Permanente Southern California (KPSC) healthcare system from 29 April to 29 July, 2022.^4^ As a comprehensive, integrated care organization, KPSC delivers healthcare across telehealth, outpatient, emergency department, and inpatient settings for over 4.7 million members. Electronic health records (EHRs) across all clinical settings, together with laboratory, pharmacy, and immunization data, provide a comprehensive view into care delivered by KPSC. These observations are augmented by insurance claims for out-of-network diagnoses, prescriptions, and procedures, enabling near-complete capture of healthcare interactions for KPSC members.

## Results

To compare severity of disease caused by BA.4/BA.5 and BA.2 infections, we monitored the frequency of healthcare interactions indicative of COVID-19 clinical progression occurring after an initial positive molecular SARS-CoV-2 test in any outpatient setting. Endpoints of interest included subsequent (≥1 day after testing) emergency department (ED) presentation or inpatient admission due to any cause, inpatient admission associated with an acute respiratory infection (ARI) diagnosis (**Table S1**), intensive care unit (ICU) admission, mechanical ventilation, and mortality. As KPSC implemented a home-based monitoring program for COVID-19 cases throughout the study period, with standardized criteria for ED referral and inpatient admission aiming to preserve hospital capacity, these endpoints provided consistent markers of disease progression within the sample followed from an initial outpatient test.^15^ We restricted our analytic sample to individuals who first tested positive in an outpatient setting to select on healthcare-seeking behavior within the study population, thus maximizing internal validity when comparing outcomes among BA.4/BA.5 and BA.2 cases. In total, 106,532 SARS-CoV-2 cases out of 148,105 diagnosed as outpatients at KPSC during the study period met eligibility criteria and were included in analyses. We excluded 18,799 patients without ≥1 year of continuous enrollment before their positive test, and 22,774 whose tests were not processed using the ThermoFisher TaqPath COVID-19 Combo Kit, which enabled lineage determination based on S gene target failure (SGTF; see **Methods**).

Within this sample, 59,556 (55.9%) and 46,976 (44.1%) cases were infected with the BA.4/BA.5 and BA.2 lineages, respectively. Whereas the weekly proportion of cases with BA.4/BA.5 infections expanded from 3.0% to 92.9% over the study period, the proportion progressing to hospital admission was stable over this period within the range of 0.2-0.5%, as compared to 0.7-1.0% during February, 2022 (**Figure 1**). Age distributions were similar among cases infected with either lineage, with 13.1% of all cases aged 0-17 years, 31.9% aged 18-39 years, 42.6% aged 40-64 years, and 12.4% aged ≥65 years (**Table 1**). Other case attributes including race/ethnicity, sex, body mass index, Charlson comorbidity index, neighborhood socioeconomic status, prior-year healthcare utilization, and receipt of vaccines targeting respiratory pathogens other than SARS-CoV-2 did not differ markedly between cases infected with the BA.4/BA.5 and BA.2 lineages.

**Table 1:** Characteristics of cases with BA.2 and BA.4/BA.5 lineage SARS-CoV-2 infection.

| Characteristic | n (%) |  |
| --- | --- | --- |
|  | BA.2 (No SGTF)<br>N=46,976 | BA.4/BA.5 (SGTF)<br>N=59,556 |
| Age<sup>1</sup> |  |  |
| 0-9 years | 3,117 (6.6) | 3,883 (6.5) |
| 10-19 years | 4,312 (9.2) | 4,755 (8.0) |
| 20-29 years | 5,146 (11.0) | 6,968 (11.7) |
| 30-39 years | 8,495 (18.1) | 11,324 (19.0) |
| 40-49 years | 8,857 (18.9) | 11,388 (19.1) |
| 50-59 years | 7,952 (16.9) | 10,016 (16.8) |
| 60-69 years | 5,542 (11.8) | 6,864 (11.5) |
| 70-79 years | 2,763 (5.9) | 3,371 (5.7) |
| ≥80 years | 792 (1.7) | 987 (1.7) |
| Sex |  |  |
| Female | 25,905 (55.1) | 32,302 (54.2) |
| Male | 21,071 (44.9) | 27,254 (45.8) |
| Race/ethnicity |  |  |
| White, non-Hispanic | 11,381 (24.2) | 12,718 (21.4) |
| Black, non-Hispanic | 3,276 (7.0) | 4,490 (7.5) |
| Hispanic | 21,145 (45.0) | 29,489 (49.5) |
| Asian | 7,503 (16.0) | 8,838 (14.1) |
| Pacific Islander | 481 (1.0) | 515 (0.9) |
| Other, mixed race, or unknown race | 3,190 (6.8) | 3,963 (6.7) |
| Neighborhood deprivation index<sup>1</sup> |  |  |
| Quintile 1 (least deprived) | 7,255 (15.4) | 7,748 (13.0) |
| Quintile 2 | 10,836 (23.1) | 12,645 (21.2) |
| Quintile 3 | 12,079 (25.7) | 15,326 (25.7) |
| Quintile 4 | 10,513 (22.4) | 14,397 (24.2) |
| Quintile 5 (most deprived) | 6,293 (13.4) | 9,440 (15.9) |
| Cigarette smoking<sup>1</sup> |  |  |
| Never smoker | 36,935 (78.6) | 46,195 (77.6) |
| Current smoker | 2,217 (4.7) | 3,079 (5.2) |
| Former smoker | 7,824 (16.7) | 10,282 (17.3) |
| Body mass index<sup>1</sup> |  |  |
| Underweight (<18.5) | 3,992 (8.5) | 4,866 (8.2) |
| Normal weight (18.5-24.9) | 11,983 (25.5) | 14,383 (24.2) |
| Overweight (25.0-29.9) | 14,203 (30.2) | 18,108 (30.4) |
| Obese (30-39.9) | 13,072 (27.8) | 17,257 (29.0) |
| Morbidly obese (≥40) | 3,726 (7.9) | 4,942 (8.3) |
| Charlson comorbidity index |  |  |
| 0 | 34,661 (73.8) | 43,825 (73.6) |
| 1-2 | 9,791 (20.8) | 12,460 (20.9) |
| 3-5 | 1,937 (4.1) | 2,510 (4.2) |
| ≥6 | 597 (1.3) | 761 (1.3) |
| Prior year outpatient visits |  |  |
| 0-4 | 13,281 (28.3) | 18,689 (31.4) |
| 5-9 | 12,924 (27.5) | 16,595 (27.9) |
| 10-14 | 7,546 (16.1) | 8,94 (15.1) |
| 15-19 | 4,375 (9.3) | 5,262 (8.8) |
| 20-29 | 4,582 (9.8) | 5,287 (8.9) |
| ≥30 | 4,268 (9.1) | 4,729 (7.9) |
| Prior year ED visits |  |  |
| 0 | 39,281 (83.6) | 49,909 (83.8) |
| 1 | 5,616 (12.0) | 7,193 (12.1) |
| 2 | 1,315 (2.8) | 1,580 (2.7) |
| ≥3 | 764 (1.6) | 874 (1.5) |
| Prior year inpatient admissions |  |  |
| 0 | 45,012 (95.8) | 57,288 (96.2) |
| 1 | 1,715 (3.7) | 1,946 (3.3) |
| 2 | 176 (0.4) | 231 (0.4) |
| ≥3 | 73 (0.2) | 91 (0.2) |
| Receipt of other vaccinations |  |  |
| 2021-22 season influenza vaccine | 26,704 (56.8) | 33,082 (55.5) |
| 13-valent pneumococcal conjugate vaccine | 10,677 (22.7) | 13,058 (21.9) |
| 23-valent pneumococcal polysaccharide vaccine | 12,477 (26.6) | 15,990 (26.8) |
| Receipt of Paxlovid<sup>2</sup> |  |  |
| Not received within 14 days of diagnosis | 44,550 (94.8) | 56,149 (94.3) |
| Received within 14 days of diagnosis | 2,426 (5.2) | 3,407 (5.7) |

SGTF: S gene target failure, here interpreted as a proxy for SARS-CoV-2 variant; CI: Confidence interval.
<sup>1</sup>Multiple imputation was used to address missing data; numbers may not add to column totals where missing values occur. The number of missing observations is as follows for the indicated covariates: age: 1 (<0.1%) BA.4/BA.5 cases, 0 (0.0%) BA.2 cases; neighborhood deprivation index: 43 (0.1%) BA.4/BA.5 cases, 38 (0.1%) BA.2 cases; cigarette smoking: 9,600 (16.1%) BA.4/BA.5 cases; 7,174 (15.3%) BA.2 cases; BMI: 10,917 (18.3%) BA.4/BA.5 cases; 7,616 (16.2%) BA.2 cases.
<sup>2</sup>Logistic regression analyses reported in **Table 2** did not include receipt of Paxlovid as a covariate predicting infecting lineage. Cox proportional hazards models reported in **Table 3**, **Table 4**, **Table S2**, and **Table S3** censored at timing of first Paxlovid dispense.

**Figure 1:**
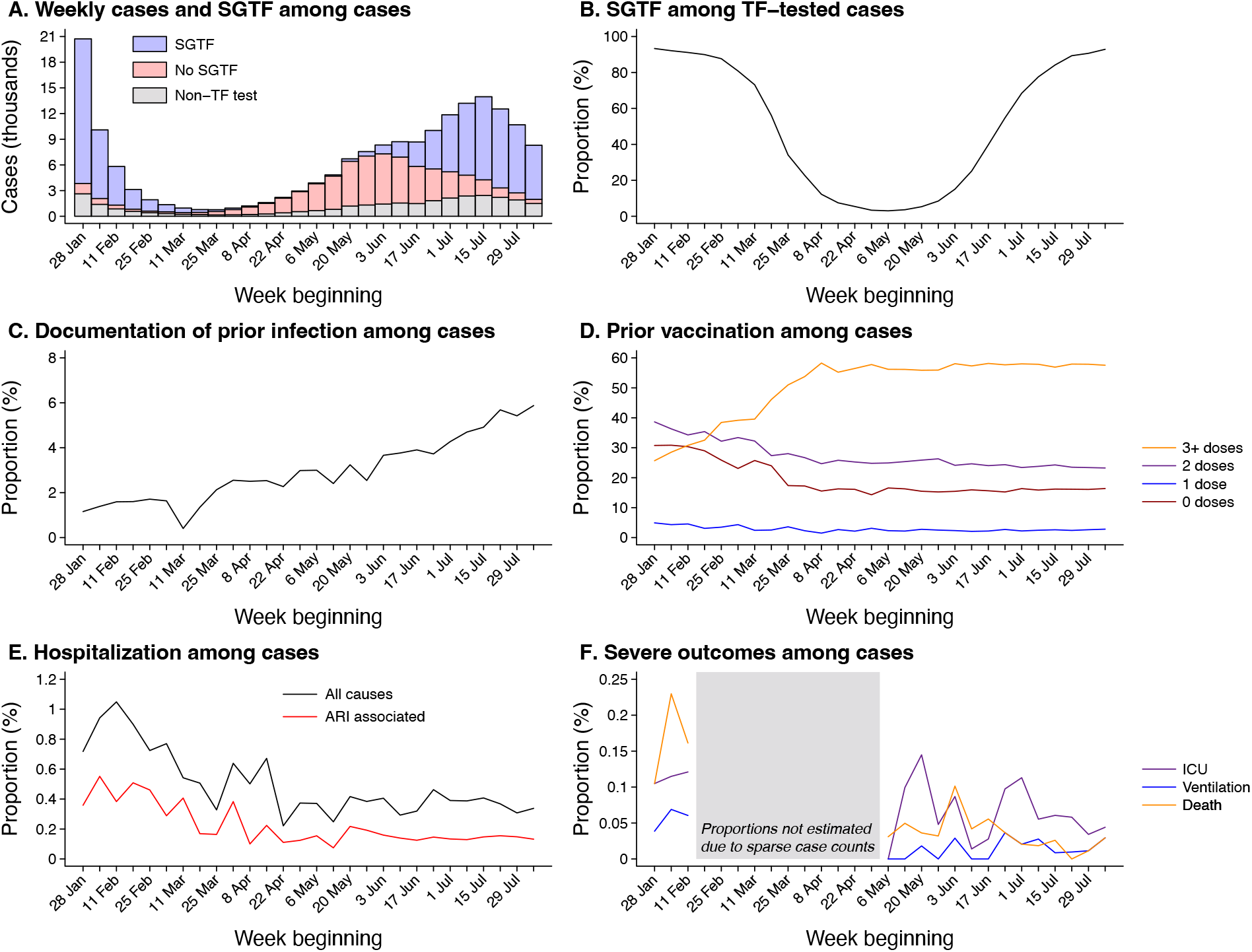
Attributes and clinical outcomes among cases diagnosed in outpatient settings. We first illustrate total outpatient-diagnosed cases, distinguishing those not tested using the TaqPath ThermoFisher COVID-19 Combo Kit (TF) assay and those determined to exhibit or not exhibit S gene target failure (SGTF; **a**). All subsequent plots are restricted to the eligible sample of outpatient cases tested using the TF assay, including the proportion of cases exhibiting SGTF (**b**); the proportion of cases with a history of prior documented infection (**c**); the proportion of cases who previously received 0, 1, 2, or ≥3 COVID-19 vaccine doses (**d**); the proportion of cases hospitalized within 30 days following their positive test (**e**), and the proportion of cases experiencing severe outcomes of intensive care unit (ICU) admission, mechanical ventilation, or death within 60 days after their positive test (**f**). The gray shaded area in panel **f** delineates weeks with <3000 cases, precluding reliable estimation of rare endpoints.

**Figure 2:**
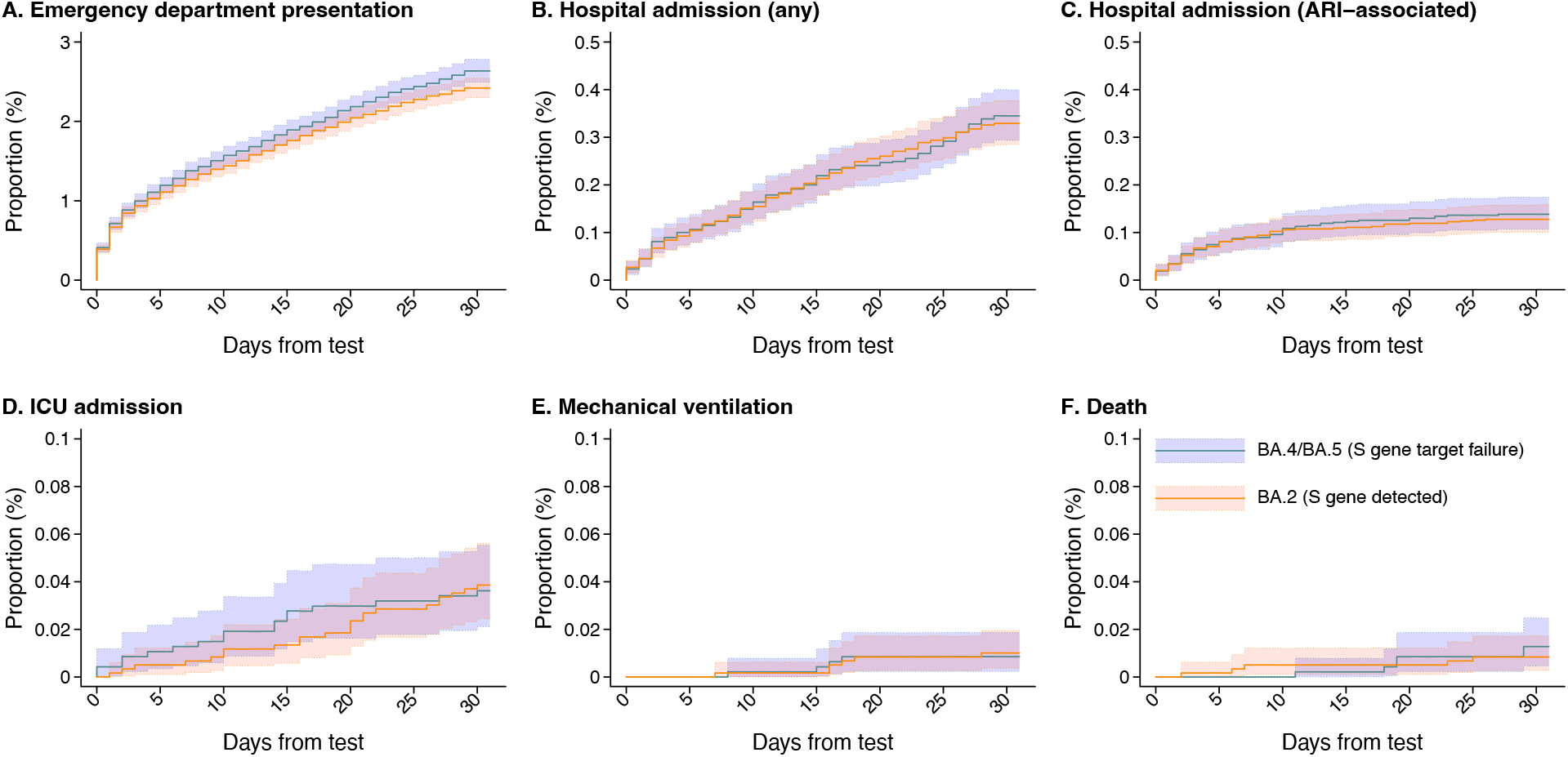
Clinical outcomes among cases with BA.2 and BA.4/BA.5 lineage SARS-CoV-2 infection, tested 29 April, 2022 to 29 July, 2022. Plots illustrate cumulative 30-day risk of severe clinical outcomes among cases first ascertained in outpatient settings, stratified by SGTF status for infecting subvariant (BA.4/BA.5 [SGTF]: orange; BA.2 [No SGTF]: blue), for endpoints of any emergency department (ED) presentation (**a**); any inpatient admission (**b**); inpatient admission associated with an acute respiratory infection (ARI) diagnosis (**c**); intensive care unit (ICU) admission (**d**); mechanical ventilation (**e**), and death (**f**). Shaded areas denote 95% confidence intervals around median estimates (center lines).

Among BA.4/BA.5 cases, 15.8% had not received any COVID-19 vaccine doses, while 2.5%, 23.6%, 48.5%, and 9.6% had received 1, 2, 3, and ≥4 doses, respectively, before their diagnosis (**Table 2**). Among BA.2 cases, 16.1%, 2.4%, 25.0%, 50.0%, and 6.6% had received 0, 1, 2, 3, and ≥4 doses, respectively. In logistic regression analyses adjusting for all measured covariates among cases, including calendar time (measured as the week or weekend of diagnosis), adjusted odds of having received 3 and ≥4 COVID-19 vaccine doses were 1.15 (95% confidence interval: 1.09-1.21) and 1.38 (1.27-1.49) fold higher among BA.4/BA.5 cases than BA.2 cases. As compared to 5.3% of BA.4/BA.5 cases, 3.1% of BA.2 cases had documentation of a prior SARS-CoV-2 infection ≥90 days before their positive test. Adjusted odds of documented prior infection were 1.55 (1.43-1.69) fold higher among BA.4/BA.5 cases than among contemporaneous BA.2 cases.

**Table 2:**
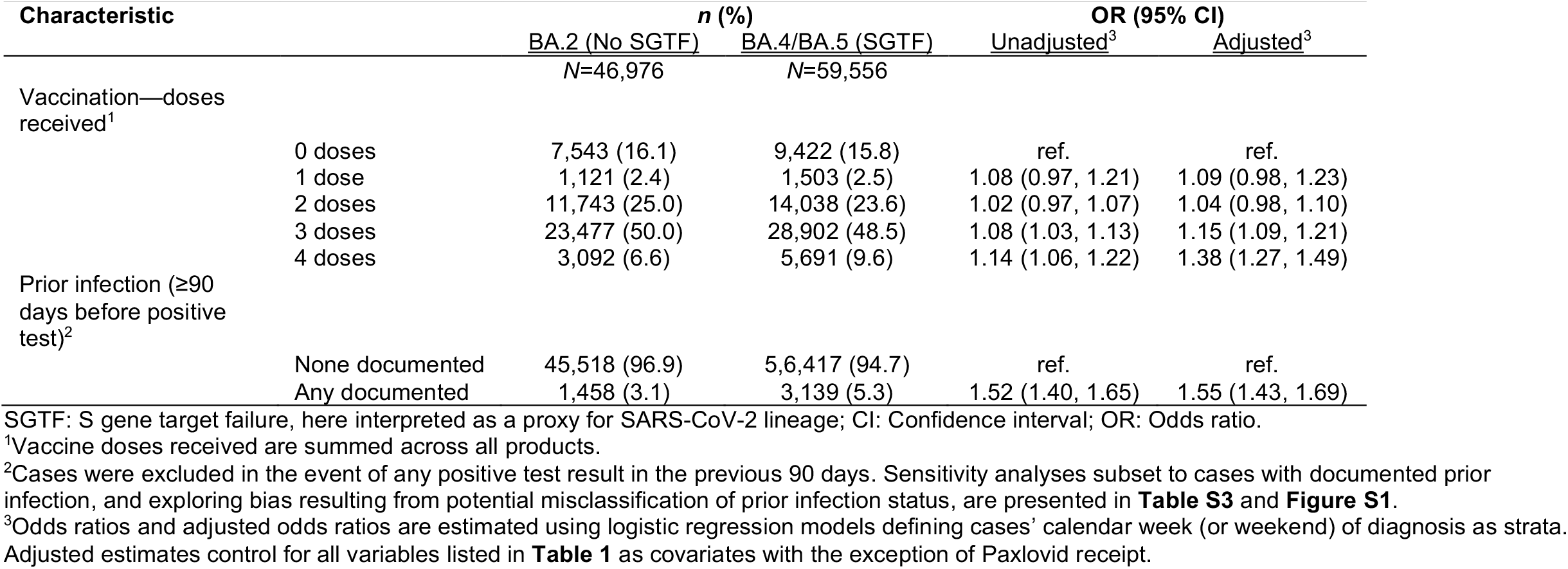
Prior vaccination and documented SARS-CoV-2 infection among cases with BA.2 and BA.4/BA.5 lineage SARS-CoV-2 infection.

Following a positive outpatient test, crude 30-day incidence of ED presentation, any inpatient admission, and inpatient admission associated with acute respiratory infection (ARI) diagnoses was 24.2, 3.3, and 1.3 per 1,000 cases with BA.4/BA.5 infection, respectively (**Figure 1**; **Table 3**). Similarly, for those with BA.2 infection, crude 30-day incidence of the same outcomes was 26.4, 3.4, and 1.4 per 1,000 cases. Higher-acuity endpoints of intensive care unit (ICU) admission, mechanical ventilation, and death occurred in a far smaller proportion of cases. Crude incidence of ICU admission, mechanical ventilation, and mortality per 10,000 cases over the first 30 days after diagnosis was 3.7, 1.0, and 0.8 among BA.4/BA.5 cases and 3.4, 0.9, and 1.3 among BA.2 cases, respectively. In analyses restricted to cases eligible for follow-up of ≥60 days, crude incidence of ICU admission, mechanical ventilation, and death was 5.3, 1.1, and 1.3 per 10,000 BA.4/BA.5 cases, and 6.8, 1.3, and 3.9 per 10,000 BA.2 cases, respectively.

**Table 3:** Association of infecting lineage with risk of severe clinical outcomes among cases tested 29 April, 2022 to 29 July, 2022.

| Clinical endpoint | Infecting lineage | *n* (%)<sup>1</sup> | Events<br>Rate per 100,000 person-days | Hazard ratio (95% CI) |  |
| --- | --- | --- | --- | --- | --- |
|  |  |  |  | Unadjusted | Adjusted |
| Emergency department presentation—30 days | BA.2 (*S gene detected*) | 1,238 (2.6) | 93.2 | ref. | ref. |
|  | BA.4/BA.5 (*S gene target failure*) | 1,441 (2.4) | 86.7 | 0.91 (0.82, 1.01) | 0.96 (0.87, 1.06) |
| Hospital admission—30 days | BA.2 (*S gene detected*) | 162 (0.34) | 11.8 | ref. | ref. |
|  | BA.4/BA.5 (*S gene target failure*) | 196 (0.33) | 11.6 | 0.92 (0.70, 1.21) | 0.96 (0.73, 1.26) |
| ARI-associated hospital admission—30 days | BA.2 (*S gene detected*) | 65 (0.13) | 4.7 | ref. | ref. |
|  | BA.4/BA.5 (*S gene target failure*) | 76 (0.13) | 4.5 | 1.02 (0.65, 1.60) | 1.09 (0.70, 1.69) |
| Intensive care unit admission—60 days | BA.2 (*S gene detected*) | 29 (0.068) | 1.2 | ref. | ref. |
|  | BA.4/BA.5 (*S gene target failure*) | 13 (0.049) | 1.1 | 0.62 (0.33, 1.17) | 0.68 (0.36, 1.27) |
| Mechanical ventilation—60 days | BA.2 (*S gene detected*) | 5 (0.012) | 0.2 | -- | -- |
|  | BA.4/BA.5 (*S gene target failure*) | 4 (0.015) | 0.3 | -- | -- |
| All-cause mortality—60 days | BA.2 (*S gene detected*) | 18 (0.068) | 0.7 | -- | -- |
|  | BA.4/BA.5 (*S gene target failure*) | 4 (0.0094) | 0.3 | -- | -- |

CI: Confidence interval. Estimates indicate the adjusted hazard ratios (aHR) of each outcome, comparing cases with BA.4/BA.5 infection to those with BA.2 infection, estimated via Cox proportional hazards models including strata for cases' week of diagnosis and all covariates listed in **Table 1** and **Table 2**.
<sup>1</sup>Proportions calculated among 46,976 BA.2 cases and 59,556 BA.4/BA.5 cases followed ≥30 days (for endpoints of emergency department presentation and hospital admission), and among 42,746 BA.2 cases and 26,339 BA.4/BA.5 cases followed ≥60 days (for endpoints of intensive care unit admission, mechanical ventilation, and all-cause mortality).

After adjustment for calendar time as well as clinical and epidemiologic factors listed in **Table 1** and **Table 2**, we did not identify independent associations of BA.4/BA.5 lineage infection with risk of any of the studied clinical outcomes (**Table 3**). Compared to observations among BA.2 cases, adjusted hazards of ED presentation and hospital admission among BA.4/BA.5 cases were 0.96 (0.87-1.06) and 0.96 (0.73-1.26) fold as high over 30 days following diagnosis. Likewise, adjusted hazards of ED presentation and hospital admission were 0.95 (0.84-1.07) and 0.96 (0.73-1.27) fold as high among BA.4/BA.5 cases as compared to BA.2 cases over the first 15 days after diagnosis (**Table S2**), a period during which such outcomes have greater specificity as markers of COVID-19 progression.^16,17^ Consistent with this observation, adjusted hazards ratios (aHRs) comparing BA.4/BA.5 cases to BA.2 cases were 1.09 (0.70-1.69) for ARI-associated hospital admission over 30 days after diagnosis, and 0.68 (0.36-1.27) for ICU admission over 60 days after diagnosis (**Table 3**). Instances of mechanical ventilation and death were too infrequent within the sample to support multivariate regression analyses adjusting for potential confounding factors.

Within these analyses, prior COVID-19 vaccination remained independently associated with protection against progression to ED presentation, hospital admission, ARI-associated hospital admission, and ICU admission for both BA.4/BA.5 and BA.2 cases (**Table 4**; **Table S3**). Effect size estimates for associations of prior COVID-19 vaccination and documented prior infection with each clinical outcome did not differ appreciably for BA.4/BA.5 cases and BA.2 cases.

**Table 4:**
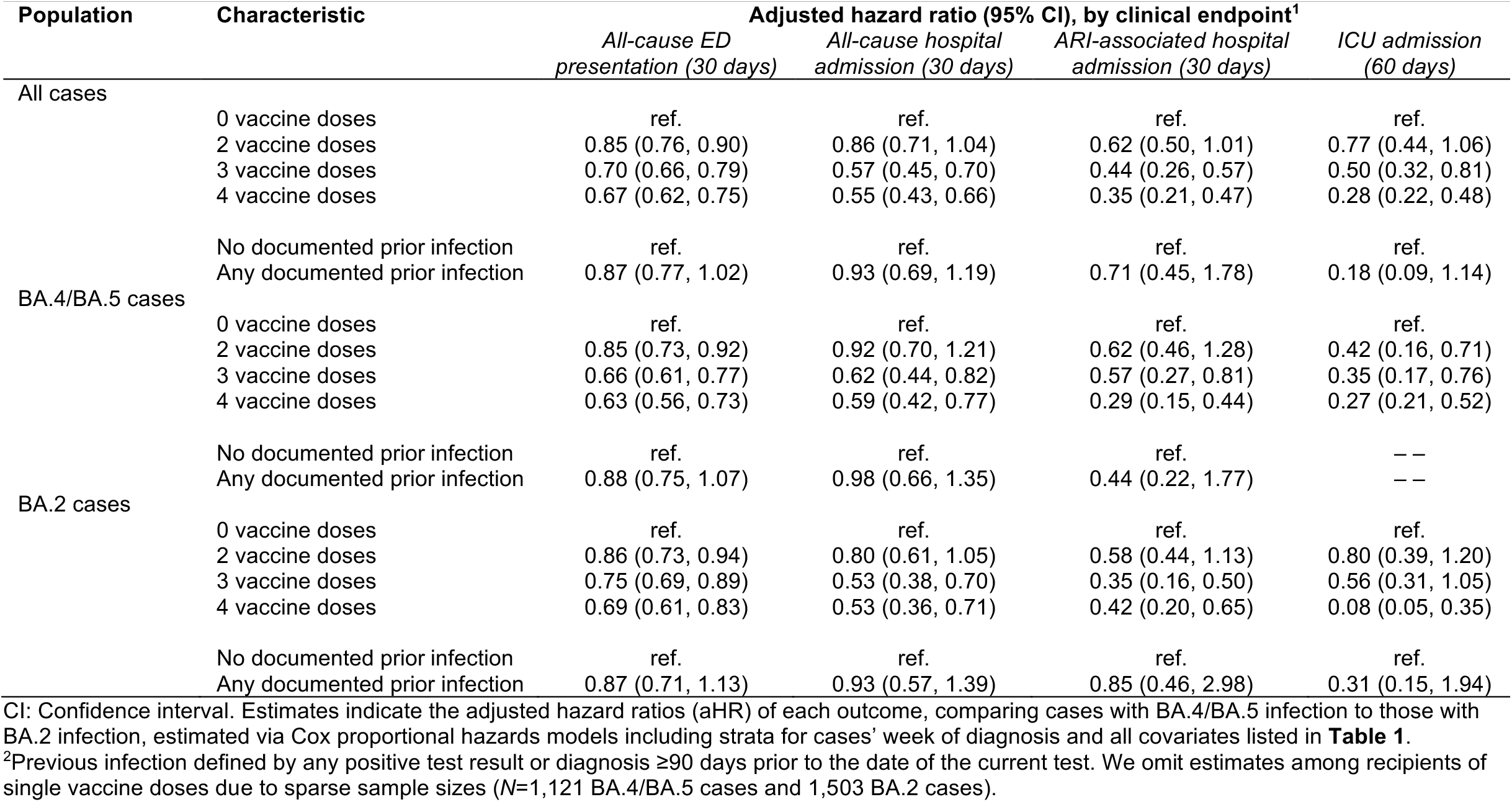
Association of prior vaccination or infection with risk of severe clinical outcomes among cases tested 29 April, 2022 to 29 July, 2022.

Because underdiagnosis of mild or asymptomatic infections could hinder our ability to control for individuals’ history of SARS-CoV-2 infection, we repeated these analyses within the subgroup of 4,597 cases (3,139 and 1,458 with BA.4/BA.5 and BA.2 infections, respectively) with documented history of SARS-CoV-2 infection ≥90 days before their index test during the study period. Within this analysis, adjusted hazards of emergency department presentation over 15 and 30 days following diagnosis were 0.81 (0.43-1.52) and 0.87 (0.54-1.41) fold as high among BA.4/BA.5 cases as among BA.2 cases, while adjusted hazards of hospital admission over 30 days were 1.45 (0.29-7.14) fold as high (**Table S4**). However, event counts were prohibitively low for adjusted analyses of ARI-associated hospital admission and higher-acuity outcomes such as ICU admission within this subgroup.

To overcome these limitations to statistical power, we further undertook risk-of-bias analyses allowing for differential degrees of under-detection of prior SARS-CoV-2 infection among individuals who ultimately experienced, or did not experience, each clinical outcome, consistent with the framework of prior analyses.^4^ Across the range of parameters considered, we did not identify scenarios where 95% confidence intervals would rule out the null hypothesis of equivalent risk of hospital admission for any cause, ARI-associated hospital admission, or ICU admission among BA.4/BA.5 and BA.2 cases (**Figure S1**). Differences in risk of ED presentation over 15 days and 30 days were expected to meet this threshold of statistical significance only when true prevalence of prior infection was modeled as ≥3-fold higher-than-observed among cases who ultimately presented to the ED, and ≥9-fold higher-than-observed among cases who did not. However, even under this scenario, bias-corrected effect sizes were expected to convey only modest differences in risk among BA.4/BA.5 and BA.2 cases (aHRs equal to 1.21 [1.07-1.37] and 1.22 [1.10-1.35], respectively, for ED presentation over 15 and 30 days among BA.4/BA.5 cases as compared to BA.2 cases).

## Discussion

Our analysis has provided insight into several characteristics of SARS-CoV-2 BA.4/BA.5 Omicron lineage infections. First, outpatient-diagnosed BA.4/BA.5 cases had 55% higher adjusted odds of a prior documented infection than time-matched BA.2 cases, as well as modestly higher adjusted odds of having received ≥3 COVID-19 vaccine doses. These findings corroborate earlier suggestions of immune escape in BA.4/BA.5 infections, which to date have been based largely on data from genomic^10^ and neutralization^11,12,18^ analyses rather than direct clinical evidence. Reassuringly, however, our findings are consistent with previous evidence that vaccination remains protective against severe disease associated with the BA.4/BA.5 lineages, at levels comparable to those reported for the BA.2 lineage.^5,19–21^ Within our large sample of 49,976 BA.2 cases and 59,556 BA.4/BA.5 cases, vaccination was not associated with statistically meaningful differences in estimates of protection against progression from an initial outpatient diagnosis to subsequent illness requiring ED presentation or either hospital or ICU admission. As our study is limited to infected cases who received clinical molecular testing, it is important to note our findings do not measure the effectiveness of prior infection or vaccination against infection with either lineage BA.4/BA.5 or BA.2 lineages. However, at least one previous study has further demonstrated that prior infection, especially with BA.1 or BA.2 Omicron lineages, remains modestly protective against BA.4/BA.5 infection, although at lower levels than those seen for earlier Omicron lineages and pre-Omicron variants.^22^

We also identify that the BA.4/BA.5 infections were not associated with enhanced risk of subsequent healthcare utilization indicative of disease progression, including ED presentation, hospital admission, or other severe endpoints, relative to BA.2 infections. As we have established in prior work that BA.2 and BA.1 lineage infections likewise do not differ in clinical severity within the KSPC population,^4^ our findings suggest that reductions in the severity of disease caused by BA.1 lineage Omicron infections, relative to the Delta variant, have persisted with BA.4/BA.5. While estimates of the severity of illness associated with BA.4/BA.5 lineage infections remain limited, our findings are consistent with those of several other studies. During the first weeks following BA.4/BA.5 emergence in South Africa, BA.4/BA.5 infections did not differ in severity from BA.1 infections, although statistical power in these analyses was constrained (*n*=1,806 BA.4/BA.5 cases analyzed) and data on cases’ clinical comorbidities and healthcare-seeking behavior were not available to fully support causal inference addressing the role of infecting variant.^9^ Consistent with this finding, risk of hospital admission during the BA.4/BA.5 and BA.1 waves in South Africa did not differ within analyses of all diagnosed cases.^23^ Whereas a population-based study in Denmark suggested moderately increased risk of hospital admission among BA.5 cases as compared to BA.2 cases,^21^ this analysis did not include adjustment for potentially relevant confounders including individuals’ healthcare-seeking behavior and calendar time. Our analyses sought to adjust for these variables based on the observed association of prior vaccination or infection with heightened risk of BA.4/BA.5 breakthrough infection, and because patient and provider demand for clinical SARS-CoV-2 testing changed markedly over the course of the BA.2 and BA.4/BA.5 waves, as public health mitigation measures were relaxed and access to home antigen testing expanded. Emergence of BA.5 was not associated with increased burden in hospital settings within Denmark, consistent with our findings and contrary to associations reported at the level of individual cases, where differing sources of confounding may apply.^24^

Our analysis has several limitations. As our sample is restricted to individuals receiving outpatient molecular testing, our findings do not convey the comprehensive burden of the BA.4/BA.5 and BA.2 lineages in the KPSC healthcare system, including cases who were admitted upon their initial presentation. Rather, this analytic framework enabled us to maximize internal validity for our primary study questions comparing BA.4/BA.5 and BA.2 cases with similar healthcare-seeking behavior, and from a similar point in their disease progression. Prior infections are likely undercounted among both BA.4/BA.5 and BA.2 cases. Because this misclassification may obscure the true magnitude of differences in prevalence of prior infection among cases acquiring each lineage, the increase in odds of prior infection among BA.4/BA.5 cases as compared to BA.2 cases likely exceeds our estimate of 55%. However, our findings of equivalent risk of severe clinical outcomes with each lineage are unlikely to be driven by this factor alone. Sensitivity analyses identified BA.4/BA.5 lineage infections would be associated with higher risk of ED presentation only under extreme scenarios where ≤1 in 9 prior infections were recorded among cases who did not require ED care (representing 97.5% of cases analyzed). Even under such a scenario, bias-corrected adjusted hazards ratio estimates were expected to identify only ~20% higher risk of ED presentation among BA.4/BA.5 cases as compared to BA.2 cases; differences in risk of other endpoints were not expected to reach statistical significance even within our large sample of 106,532 cases. While the true prevalence of unascertained prior infection in this population is not precisely known, this scenario represents a considerable departure from prior estimates of the reporting fraction in California.^25^ It is important to note that our analyses do not distinguish causes of ED presentations and hospital admission, although ED presentations and hospital admissions occurring within 15 days of cases’ first positive test, and ARI-associated hospital admissions, likely have greater specificity for indicating healthcare interactions precipitated by COVID-19 illness.^16,17^ Last, our analyses do not distinguish cases infected with BA.4 and BA.5, or cases with BA.2.12.1 versus other BA.2 sublineages, which may be associated with distinct epidemiologic and clinical characteristics.

While it is encouraging that we find BA.4/BA.5 lineage infections are not associated with differential severity in comparison to other Omicron lineages, it is important to note that disease burden is influenced by further variant-specific properties including the intrinsic capacity to transmit and to infect individuals with immunity from prior vaccination or infection.^26^ These fitness advantages are relevant to consider in the context of BA.4/BA.5, which outcompeted BA.2 in the context of substantial population immunity.^10^ As of the week ending 29 October, BA.5 continues to account for over half of newly-diagnosed SARS-CoV-2 cases in the US, although other lineages including BQ.1 and BF.7 are identified in an increasing proportion of cases. As novel SARS-CoV-2 variants continue to emerge, monitoring associations of novel circulating lineages with risk of severe illness and post-vaccination breakthrough infection will provide key insight to inform public health responses.

## Data Availability

Individual level clinical data are not publicly available for sharing.

## Acknowledgments

This work was funded by the US Centers for Disease Control and Prevention (CDC; grant 75D3-121C11520 to SYT). The findings and conclusions in this report are those of the authors and do not necessarily represent the official position of the CDC. JAL was supported by grant R01-AI14812701A1 from the National Institute for Allergy and Infectious Diseases (US National Institutes of Health), which had no role in design or conduct of the study, or the decision to submit for publication.

## Author contributions statement

JAL, VH, and SYT contributed to the study concept and design. JAL, VH, and SYT led acquisition and statistical analysis of data. JAL and SYT led interpretation of data. JAL drafted the manuscript, and VH and SYT critically revised the manuscript for important intellectual content. SYT obtained funding and provided supervision.

## Methods

### Setting, procedures, and study population

Care delivery within KPSC has been described previously.^4^ Briefly, approximately 19% of the population of southern California receives care from KPSC through employer-provided, pre-paid, or federally sponsored insurance plans. In-network care delivery data encompassing diagnoses (and accompanying clinical notes), immunizations, laboratory tests administered and test results, and prescriptions are captured in near-real time via patient EHRs, while out-of-network care is captured through insurance claim reimbursements. Delivery of COVID-19 vaccine doses by other providers was identified via linkage to California Immunization Registry data and other health systems using the Epic EHR system. Online portals provide an automated platform for individuals to upload or notify providers of positive at-home test results or test results received from other providers.

Molecular diagnostic testing for COVID-19 was made available to all individuals receiving outpatient care from KSPC for any indication during the study period, and was used to screen all patients for SARS-CoV-2 infection at the point of hospital admission for any cause. Consistent with prior analyses,^4^ we restricted our analytic sample to cases with a positive SARS-CoV-2 test result from testing undertaken in outpatient settings using the ThermoFisher TaqPath COVID-19 Combo Kit (the most commonly used assay for outpatient testing at KPSC during the study period), with ≥1 year of continuous enrollment in KPSC health plans prior to their test date. In addition to enabling longitudinal follow-up for severe endpoint ascertainment, restricting analyses to cases tested as outpatients was expected to provide two design advantages helping to mitigate bias. First, excluding individuals first ascertained in hospital settings helped to reduce bias driven by differential healthcare-seeking behavior among cases tested as outpatients versus those who deferred testing to more severe stages of illness. Second, this approach enabled us to minimize the inclusion of cases hospitalized for other cuases who were identified incidentally via SARS-CoV-2 infection screening at admission. Although ED presentations, hospital admissions, and other study endpoints are generally rare events, individuals’ risk of each endpoint is greatly increased by SARS-CoV-2 infection. Thus, a majority of observed severe endpoints observed within the outpatient-diagnosed sample were expected to be attributable to COVID-19.

Laboratory data included qualitative (presence/absence) detection of RNA for probes targeting the SARS-CoV-2 S, nucleocapsid (N), and Orf1a/b genes. As BA.4/BA.5 lineages harbor the Δ69-70 amino acid deletion in the S protein, SGTF has been proposed elsewhere as a proxy for distinguishing BA.4/BA.5 from BA.2 lineages.^9,10^ Analyses of a randomly selected subset of SARS-CoV-2 specimens submitted for whole-genome sequencing supported this approach, with the S gene target identified in 97.7% (2,600/2,660) of BA.2 specimens and absent from 99.4% (347/349) of BA.4 specimens and 97.5% (977/1,002) of BA.5 specimens.

To ensure our analyses captured new-onset infections, we excluded cases with a positive SARS-CoV-2 test result within the prior 90 days. The study protocol was approved by the KPSC Institutional Review Board.

### Outcomes

Outcomes of interest to our analyses included: (1) any ED presentation; (2) any inpatient admission; (3) ARI-associated inpatient admission, at which physicians recorded ≥1 of the ARI diagnostic codes indicated in **Table S1**; (4) ICU admission; (5) mechanical ventilation; and (6) death. We limited follow-up time for ED presentation and hospital admission to 30 days following the initial positive outpatient test; we included follow-up time through 60 days from the initial positive outpatient test for endpoints of ICU admission, mechanical ventilation, and death, owing to the longer course of disease expected to precede such outcomes. We censored observations at study end date or at disenrollment for cases who had not experienced each outcome. As cases diagnosed in outpatient settings were enrolled in a home-based monitoring program with standardized criteria for ED referral and inpatient admission,^15^ we expected severity of illness associated with each endpoint to be internally comparable within the study cohort. Last, to facilitate our ability to measure intrinsic associations of infecting lineage with risk of progression to severe outcomes, we censored observations at dates of Paxlovid dispense for individuals who received this treatment (5.5% of patients analyzed [*n*=5,833]). Real-world effectiveness of Paxlovid in preventing adverse clinical outcomes within this population has been described elsewhere.^17,27^

### Case characteristics

We recorded the following characteristics for each case: age (defined in 10-year age bands), sex, race/ethnicity (white, black, Hispanic of any race, Asian, Pacific Islander, and other/mixed/unknown race), neighborhood deprivation index, measured at the Census block level; smoking status (current, former, or never smoker); body mass index (BMI; underweight, normal weight, overweight, obese, and morbidly obese); Charlson comorbidity index (0, 1-2, 3-5, and ≥6); prior-year emergency department visits and inpatient admissions (each defined as 0, 1, 2, or ≥3 events); prior-year outpatient visits (0-4, 5-9, 10-14, 15-19, 20-29, or ≥30 events); documented prior SARS-CoV-2 infection; and history of COVID-19 vaccination (receipt of 0, 1, 2, 3, or ≥4 doses, and time from receipt of each dose to each case’s testing date), and receipt of Paxlovid ≤14 days after the initial outpatient diagnosis date.

### Multiple imputation of missing data

Variables with missing data included cases’ age (*n*=1; 0.00094% of 106,532 cases), neighborhood deprivation index (*n*=81; 0.076% of cases), BMI (*n*=18,533; 17.4% of observations), and cigarette smoking status (*n*=16,774; 15.7% of observations). We populated 10 complete pseudo-datasets sampling from the distribution of missing values, according to the joint distribution of all measured variables, via multiple imputation, and repeated all statistical analyses across each pseudo-dataset. We pooled resulting estimates according to Rubin’s rules.^28^

### Logistic regression analysis

We compared the distributions of prior vaccination status and prior infection status among BA.4/BA.5 cases versus BA.2 cases via logistic regression. Models controlled for all variables listed above, with the exception of Paxlovid receipt (which occurred after diagnosis), to define aORs in relation to infecting lineage. Models included distinct intercepts for each calendar week to control for potential changes in testing and healthcare-seeking practices over the period of BA.4/BA.5 emergence.

### Survival analysis

We fit Cox proportional regression models including data from all outpatient-diagnosed cases, censoring at either the study end date, end of follow-up, disenrollment, or date of Paxlovid dispense. Models defined covariates for each case characteristic listed above. Models defined strata according to cases’ calendar week of testing to control for potential changes in testing and healthcare-seeking practices over the period of BA.4/BA.5 emergence. We verified the proportional hazards assumption for all models by testing for non-zero slopes of the Schoenfeld residuals.^29,30^

### Sensitivity analyses

Because protection from prior infection could contribute to lower risk of clinical progression among a higher proportion of cases with BA.4/BA.5 infection than BA.2 infection,^24^ we undertook several sensitivity analyses aiming to determine whether our results were robust to bias driven by potentially differential prevalence of unrecorded prior infections among cases infected with each lineage. First, we repeated our primary survival analyses within the subset of cases known to have experienced a prior infection, as differential prevalence of prior infection could not lead to differences in disease progression within this stratum. However, sample sizes were inadequate to allow similar analyses for all endpoints within this subset. We therefore conducted risk-of-bias analyses allowing for non-differential or differential undercounting of prior infections among cases with BA.4/BA.5 and BA.2 infection, similar to prior work in the study population^4^ and described in detail below.

Within each imputed pseudo-dataset, we fit logistic regression models to define cases’ propensity for prior SARS-CoV-2 infection as a function of all measured characteristics (including SGTF status) as well as the observed occurrence of endpoints of symptoms potentially associated with SARS-CoV-2,^16^ ED presentation, hospital admission, ICU admission, mechanical ventilation, and death. To account for potentially higher-than-observed prevalence of prior infection among all cases, we repeated analyses resampling individual infection histories at random under an assumption that true prevalence of prior infection was *ρ* ∈ (1, 1.5, 2, 3) times higher than that observed based on fitted propensity scores. To further allow for potentially higher prevalence of prior infection among cases who were protected from experiencing clinical outcomes, we multiplied the estimated propensity of prior infection within these groups by a factor equal to *α* × ρ, for *α* ∈ (1, 1.5, 2, 3), thus allowing up to 9-fold higher-than-observed prevalence of prior infection among individuals who were protected from experiencing each endpoint. We plot resulting estimates of the bias-corrected adjusted hazards ratios of each outcome, comparing BA.4/BA.5 cases to BA.2 cases, in **Figure S1**.

## Software

We conducted analyses using R version 4.0.3 (R Foundation for Statistical Computing, Vienna, Austria). We used the survival^31^ package for time-to-event analyses, and the Amelia II package^32^ for multiple imputation.

## Data availability

Individual-level data reported in this study are not publicly shared.

## Code availability

Analysis code will be made available from github.com/joelewnard/ba4ba5severity upon publication.

**Table S1:**
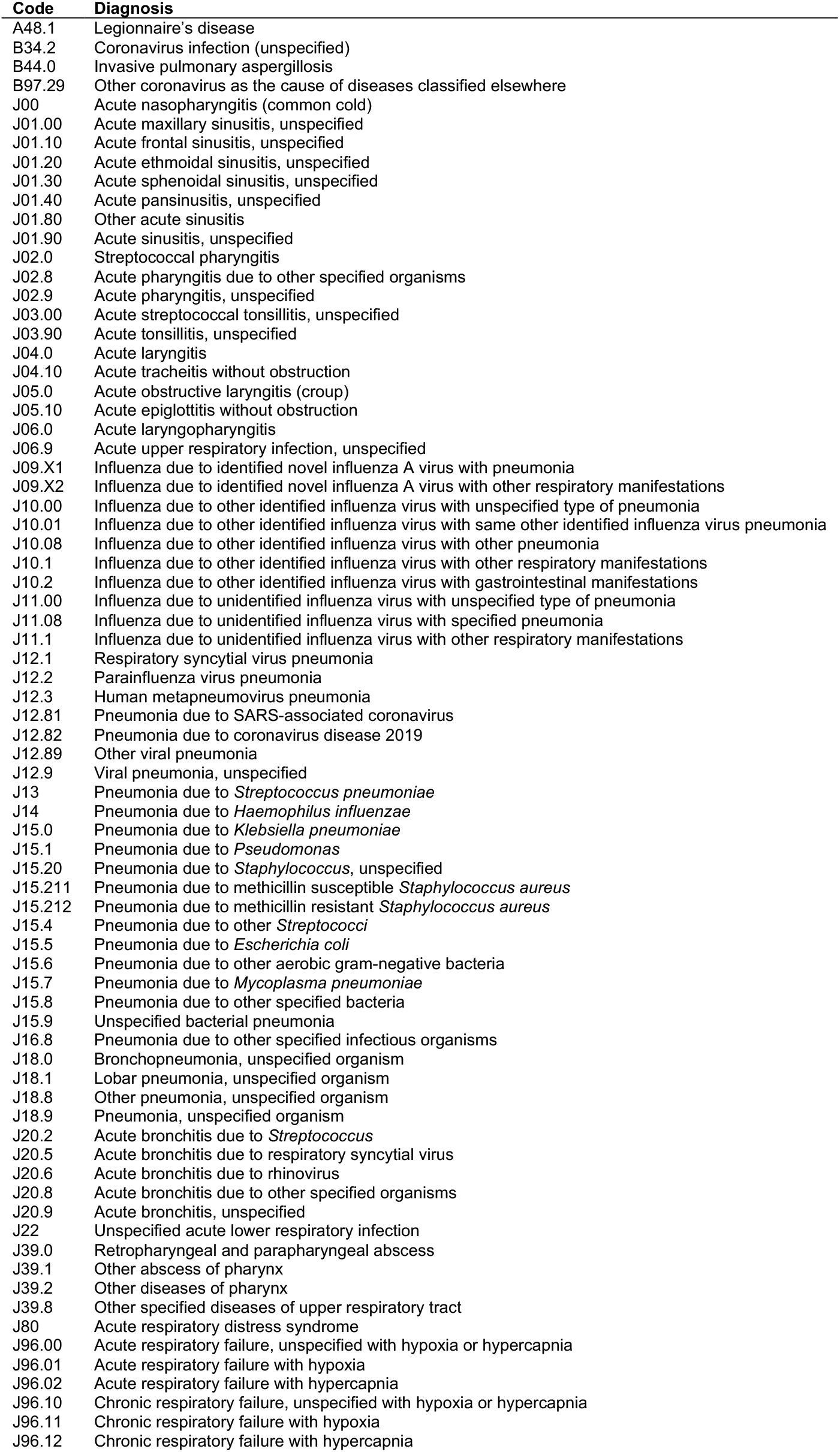
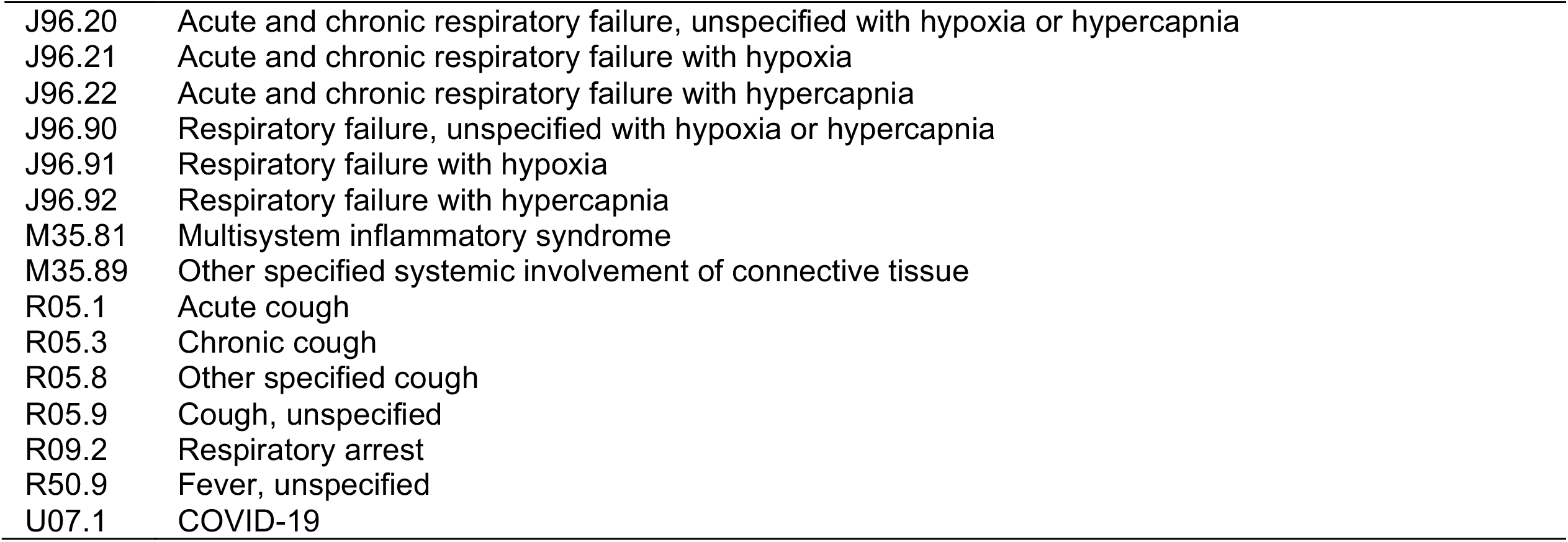
Diagnosis codes used to identify acute respiratory infection-associated hospital admissions.

| Code | Diagnosis |
| --- | --- |
| A48.1 | Legionnaire's disease |
| B34.2 | Coronavirus infection (unspecified) |
| B44.0 | Invasive pulmonary aspergillosis |
| B97.29 | Other coronavirus as the cause of diseases classified elsewhere |
| J00 | Acute nasopharyngitis (common cold) |
| J01.00 | Acute maxillary sinusitis, unspecified |
| J01.10 | Acute frontal sinusitis, unspecified |
| J01.20 | Acute ethmoidal sinusitis, unspecified |
| J01.30 | Acute sphenoidal sinusitis, unspecified |
| J01.40 | Acute pansinusitis, unspecified |
| J01.80 | Other acute sinusitis |
| J01.90 | Acute sinusitis, unspecified |
| J02.0 | Streptococcal pharyngitis |
| J02.8 | Acute pharyngitis due to other specified organisms |
| J02.9 | Acute pharyngitis, unspecified |
| J03.00 | Acute streptococcal tonsillitis, unspecified |
| J03.90 | Acute tonsillitis, unspecified |
| J04.0 | Acute laryngitis |
| J04.10 | Acute tracheitis without obstruction |
| J05.0 | Acute obstructive laryngitis (croup) |
| J05.10 | Acute epiglottitis without obstruction |
| J06.0 | Acute laryngopharyngitis |
| J06.9 | Acute upper respiratory infection, unspecified |
| J09.X1 | Influenza due to identified novel influenza A virus with pneumonia |
| J09.X2 | Influenza due to identified novel influenza A virus with other respiratory manifestations |
| J10.00 | Influenza due to other identified influenza virus with unspecified type of pneumonia |
| J10.01 | Influenza due to other identified influenza virus with same other identified influenza virus pneumonia |
| J10.08 | Influenza due to other identified influenza virus with other pneumonia |
| J10.1 | Influenza due to other identified influenza virus with other respiratory manifestations |
| J10.2 | Influenza due to other identified influenza virus with gastrointestinal manifestations |
| J11.00 | Influenza due to unidentified influenza virus with unspecified type of pneumonia |
| J11.08 | Influenza due to unidentified influenza virus with specified pneumonia |
| J11.1 | Influenza due to unidentified influenza virus with other respiratory manifestations |
| J12.1 | Respiratory syncytial virus pneumonia |
| J12.2 | Parainfluenza virus pneumonia |
| J12.3 | Human metapneumovirus pneumonia |
| J12.81 | Pneumonia due to SARS-associated coronavirus |
| J12.82 | Pneumonia due to coronavirus disease 2019 |
| J12.89 | Other viral pneumonia |
| J12.9 | Viral pneumonia, unspecified |
| J13 | Pneumonia due to *Streptococcus pneumoniae* |
| J14 | Pneumonia due to *Haemophilus influenzae* |
| J15.0 | Pneumonia due to *Klebsiella pneumoniae* |
| J15.1 | Pneumonia due to *Pseudomonas* |
| J15.20 | Pneumonia due to *Staphylococcus*, unspecified |
| J15.211 | Pneumonia due to methicillin susceptible *Staphylococcus aureus* |
| J15.212 | Pneumonia due to methicillin resistant *Staphylococcus aureus* |
| J15.4 | Pneumonia due to other *Streptococci* |
| J15.5 | Pneumonia due to *Escherichia coli* |
| J15.6 | Pneumonia due to other aerobic gram-negative bacteria |
| J15.7 | Pneumonia due to *Mycoplasma pneumoniae* |
| J15.8 | Pneumonia due to other specified bacteria |
| J15.9 | Unspecified bacterial pneumonia |
| J16.8 | Pneumonia due to other specified infectious organisms |
| J18.0 | Bronchopneumonia, unspecified organism |
| J18.1 | Lobar pneumonia, unspecified organism |
| J18.8 | Other pneumonia, unspecified organism |
| J18.9 | Pneumonia, unspecified organism |
| J20.2 | Acute bronchitis due to *Streptococcus* |
| J20.5 | Acute bronchitis due to respiratory syncytial virus |
| J20.6 | Acute bronchitis due to rhinovirus |
| J20.8 | Acute bronchitis due to other specified organisms |
| J20.9 | Acute bronchitis, unspecified |
| J22 | Unspecified acute lower respiratory infection |
| J39.0 | Retropharyngeal and parapharyngeal abscess |
| J39.1 | Other abscess of pharynx |
| J39.2 | Other diseases of pharynx |
| J39.8 | Other specified diseases of upper respiratory tract |
| J80 | Acute respiratory distress syndrome |
| J96.00 | Acute respiratory failure, unspecified with hypoxia or hypercapnia |
| J96.01 | Acute respiratory failure with hypoxia |
| J96.02 | Acute respiratory failure with hypercapnia |
| J96.10 | Chronic respiratory failure, unspecified with hypoxia or hypercapnia |
| J96.11 | Chronic respiratory failure with hypoxia |
| J96.12 | Chronic respiratory failure with hypercapnia |
| J96.20 | Acute and chronic respiratory failure, unspecified with hypoxia or hypercapnia |
| J96.21 | Acute and chronic respiratory failure with hypoxia |
| J96.22 | Acute and chronic respiratory failure with hypercapnia |
| J96.90 | Respiratory failure, unspecified with hypoxia or hypercapnia |
| J96.91 | Respiratory failure with hypoxia |
| J96.92 | Respiratory failure with hypercapnia |
| M35.81 | Multisystem inflammatory syndrome |
| M35.89 | Other specified systemic involvement of connective tissue |
| R05.1 | Acute cough |
| R05.3 | Chronic cough |
| R05.8 | Other specified cough |
| R05.9 | Cough, unspecified |
| R09.2 | Respiratory arrest |
| R50.9 | Fever, unspecified |
| U07.1 | COVID-19 |

**Table S2:**
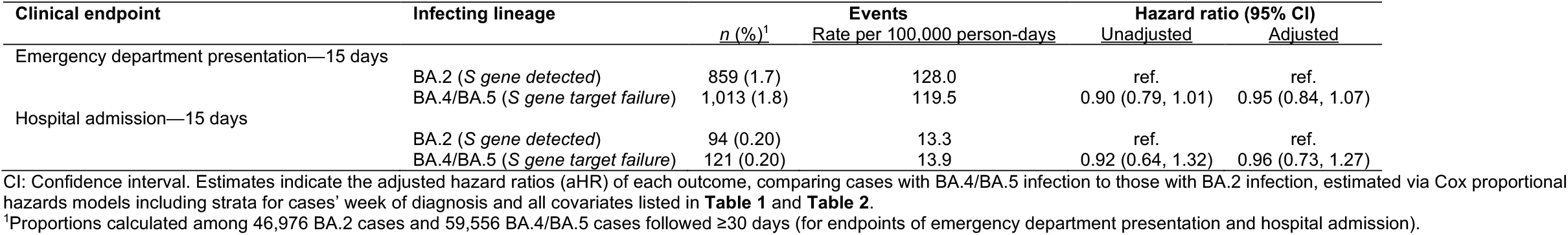
Association of infecting lineage risk of severe clinical outcomes over 15 days after diagnosis among cases tested 29 April, 2022 to 29 July, 2022.

| Clinical endpoint | Infecting lineage | Events |  | Hazard ratio (95% CI) |  |
| --- | --- | --- | --- | --- | --- |
|  |  | *n* (%)<sup>1</sup> | Rate per 100,000 person-days | Unadjusted | Adjusted |
| Emergency department presentation—15 days | BA.2 (*S gene detected*) | 859 (1.7) | 128.0 | ref. | ref. |
|  | BA.4/BA.5 (*S gene target failure*) | 1,013 (1.8) | 119.5 | 0.90 (0.79, 1.01) | 0.95 (0.84, 1.07) |
| Hospital admission—15 days | BA.2 (*S gene detected*) | 94 (0.20) | 13.3 | ref. | ref. |
|  | BA.4/BA.5 (*S gene target failure*) | 121 (0.20) | 13.9 | 0.92 (0.64, 1.32) | 0.96 (0.73, 1.27) |

CI: Confidence interval. Estimates indicate the adjusted hazard ratios (aHR) of each outcome, comparing cases with BA.4/BA.5 infection to those with BA.2 infection, estimated via Cox proportional hazards models including strata for cases' week of diagnosis and all covariates listed in **Table 1** and **Table 2**.
<sup>1</sup>Proportions calculated among 46,976 BA.2 cases and 59,556 BA.4/BA.5 cases followed ≥30 days (for endpoints of emergency department presentation and hospital admission).

**Table S3:**
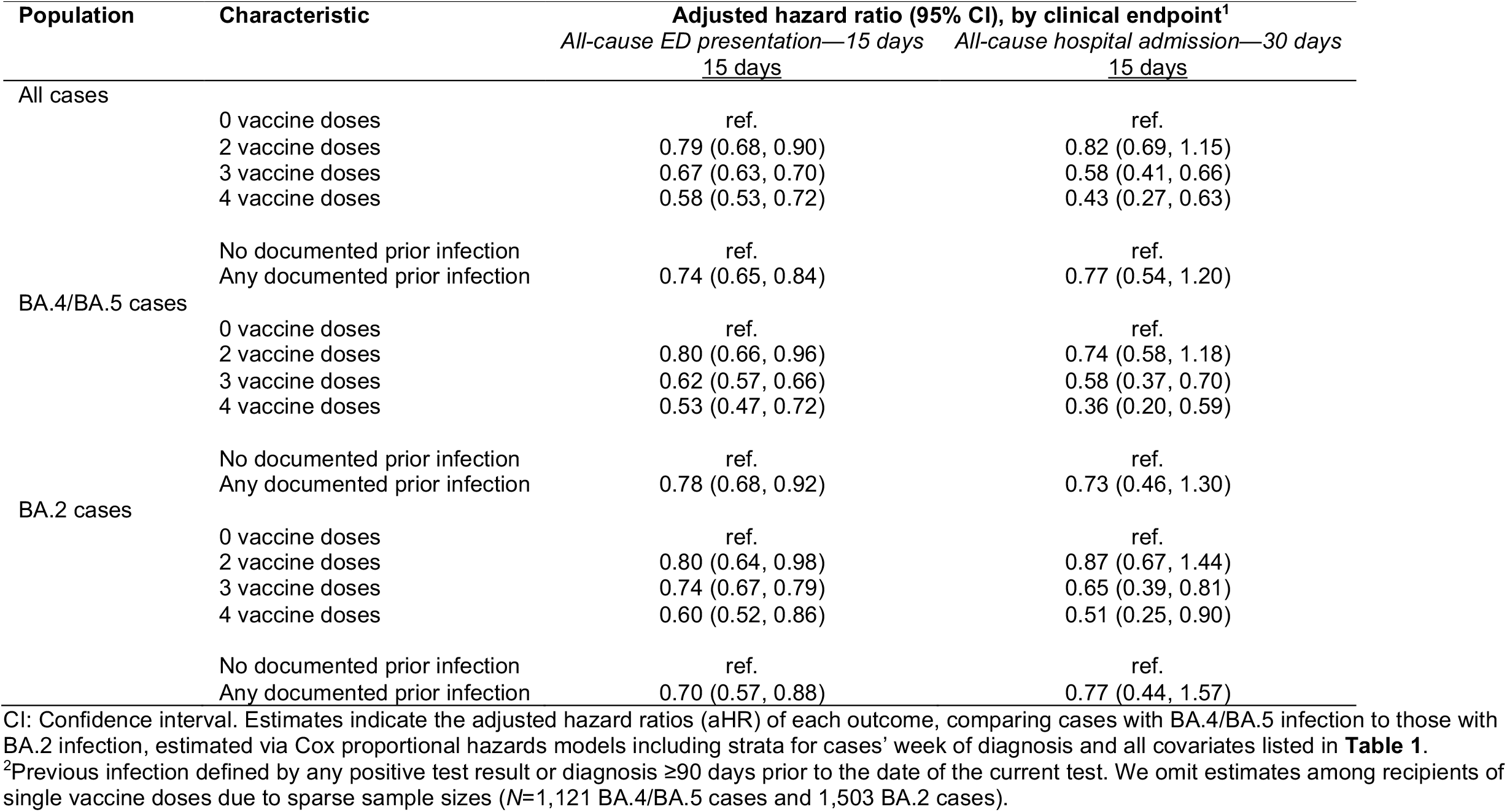
Association of prior vaccination or infection with risk of severe clinical outcomes over 15 days after diagnosis among cases tested 29 April, 2022 to 29 July, 2022.

**Table S4:** Clinical outcomes among cases with BA.2 and BA.4/BA.5 lineage SARS-CoV-2 infection with documented history of SARS-CoV-2 infection.

| Clinical outcome | Population | Events, *n* (%)<sup>1</sup> | Rate per 100,000 person-days | HR (95% CI) | aHR (95% CI) |
| --- | --- | --- | --- | --- | --- |
| Emergency department presentation—15 days | BA.2 (*S gene detected*) | 26 (1.8) | 122.9 | ref. | ref. |
|  | BA.4/BA.5 (*S gene target failure*) | 54 (1.7) | 118.8 | 0.80 (0.44, 1.48) | 0.81 (0.43, 1.52) |
| Emergency department presentation—30 days | BA.2 (*S gene detected*) | 49 (3.4) | 117.1 | ref. | ref. |
|  | BA.4/BA.5 (*S gene target failure*) | 89 (2.8) | 100.0 | 0.85 (0.54, 1.35) | 0.87 (0.54, 1.41) |
| Hospital admission—15 days | BA.2 (*S gene detected*) | 4 (0.3) | 18.1 | ref. | — |
|  | BA.4/BA.5 (*S gene target failure*) | 6 (0.2) | 13.0 | 0.80 (0.13, 5.06) | — |
| Hospital admission—30 days | BA.2 (*S gene detected*) | 7 (0.5) | 16.2 | ref. | ref. |
|  | BA.4/BA.5 (*S gene target failure*) | 11 (0.4) | 12.2 | 1.30 (0.33, 5.09) | 1.45 (0.29, 7.14) |
| ARI-associated hospital admission—30 days | BA.2 (*S gene detected*) | 4 (0.3) | 9.3 | ref. | — |
|  | BA.4/BA.5 (*S gene target failure*) | 3 (0.1) | 3.3 | 0.49 (0.06, 4.39) | — |

CI: Confidence interval. Estimates indicate the hazard ratio (HR) adjusted hazard ratio (aHR) of each outcome, comparing cases with BA.4/BA.5 infection to those with BA.2 infection, estimated via Cox proportional hazards models including strata for cases' calendar week (or weekend) of diagnosis and all covariates listed in **Table 1** and **Table 2**. Analyses are subset to cases with documented SARS-CoV-2 infection based on a positive molecular test result $\geq 90$ days before the date of the current test (3,139 BA.4/BA.5 cases and 1,458 BA.2 cases). We omit analyses of endpoints of ICU admission, mechanical ventilation, and death due to sparse counts.

**Figure S1:**
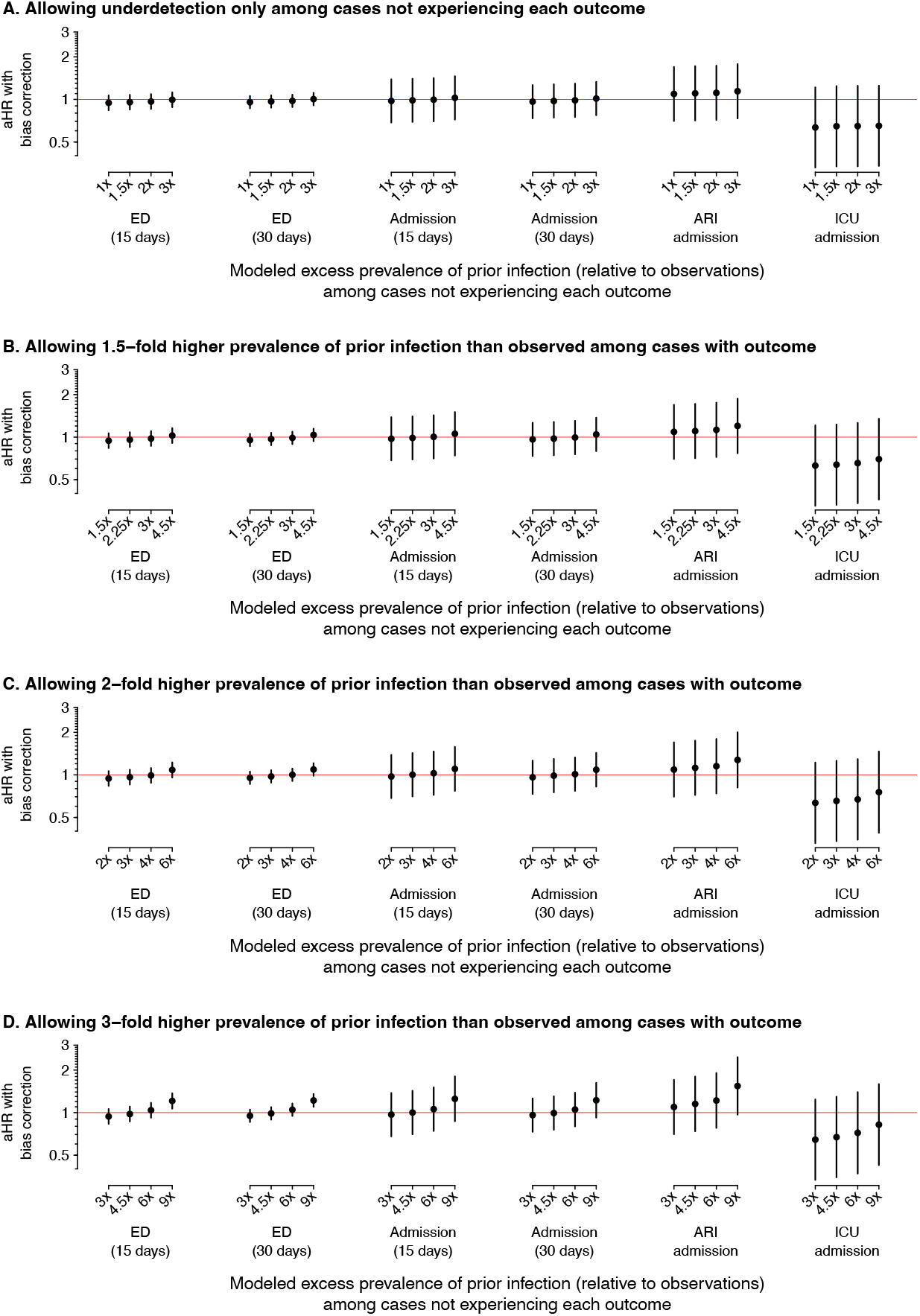
We present estimates of the bias-corrected adjusted hazards ratio (aHR), together with 95% confidence intervals (CIs; vertical lines), under various scenarios with respect to the proportion of prior infections observed among individuals who experienced each outcome (*ρ*^−1^, for *ρ*∈ {1, 1.5, 2, 3}) and among individuals who did not experience each outcome (*α*^−1^, for *α* ∈ {1, 1.5, 2, 3}). Individuals’ baseline propensity for having experienced prior infection is estimated by logistic regression, thus accounting for risk factors including vaccination and the individual’s infection with either the BA.4/BA.5 or BA.2 lineages. Panels **a, b, c, d** correspond to scenarios with *ρ*= 1, 1.5, 2, 3, respectively, among individuals who experienced each outcome, while estimates under differing values of *α* are presented side-by-side within each panel, for each endpoint. Row labels denote the product *α* × *ρ*, indicating the modeled ratio of true-to-observed infections among individuals who did not experience each clinical endpoint of interest. Statistically-significant bias-corrected aHR estimates (signified by 95% CIs that exclude the null hypothesis of no difference in risk of each outcome for cases with BA.4/BA.5 or BA.2 lineage infections) emerge only under the scenario of *ρ*= 3 and *α* = 3, for 15- and 30-day risk of ED presentation. Following bias correction, we estimate that the aHR or ED presentation comparing BA.4/BA.5 to BA.2 cases is 1.21 (95% CI: 1.07-1.37) over 15 days and 1.22 (1.10-1.35) over 30 days under this scenario.

